# Presence of SARS-CoV-2-reactive T cells in COVID-19 patients and healthy donors

**DOI:** 10.1101/2020.04.17.20061440

**Authors:** Julian Braun, Lucie Loyal, Marco Frentsch, Daniel Wendisch, Philipp Georg, Florian Kurth, Stefan Hippenstiel, Manuela Dingeldey, Beate Kruse, Florent Fauchere, Emre Baysal, Maike Mangold, Larissa Henze, Roland Lauster, Marcus A. Mall, Kirsten Beyer, Jobst Röhmel, Jürgen Schmitz, Stefan Miltenyi, Ilja Demuth, Marcel A. Müller, Martin Witzenrath, Norbert Suttorp, Florian Kern, Ulf Reimer, Holger Wenschuh, Christian Drosten, Victor M. Corman, Claudia Giesecke-Thiel, Leif Erik Sander, Andreas Thiel

**Affiliations:** Si-M / “Der Simulierte Mensch” a science framework of Technische Universität Berlin and Charité - Universitätsmedizin Berlin, Berlin, Germany; Regenerative Immunology and Aging, BIH Center for Regenerative Therapies, Charité Universitätsmedizin Berlin, Berlin, Germany; Department of Hematology, Oncology and Tumor Immunology, Charité - Universitätsmedizin Berlin; Department of Infectious Diseases and Respiratory Medicine, Charité - Universitätsmedizin Berlin; Medical Biotechnology, Institute for Biotechnology, Technische Universität Berlin; Department of Pediatric Pulmonology, Immunology and Critical Care Medicine, Charité - Universitätsmedizin Berlin; Miltenyi Biotec GmbH, Bergisch-Gladbach, Germany; Interdisciplinary Metabolism Center, Biology of Aging (BoA)-group, Charité Universitätsmedizin Berlin; Institute of Virology, Charité - Universitätsmedizin Berlin, Berlin, Germany; JPT Peptide Technologies GmbH, Berlin, Germany; Brighton and Sussex Medical School, Department of Clinical and Experimental Medicine, Brighton, UK; Berlin Institute of Health (BIH), Berlin, Germany; Max Planck Institute for Molecular Genetics, Berlin, Germany

## Abstract

Severe acute respiratory syndrome coronavirus 2 (SARS-CoV-2) has caused a rapidly unfolding pandemic, overwhelming health care systems worldwide^1^. Clinical manifestations of Coronavirus-disease 2019 (COVID-19) vary broadly, ranging from asymptomatic infection to acute respiratory failure and death^2^, yet the underlying mechanisms for this high variability are still unknown. Similarly, the role of host immune responses in viral clearance of COVID-19 remains unresolved. For SARS-CoV (2002/03), however, it has been reported that CD4^+^ T cell responses correlated with positive outcomes^3,4^, whereas T cell immune responses to SARS-CoV-2 have not yet been characterized. Here, we describe an assay that allows direct detection and characterization of SARS-CoV-2 spike glycoprotein (S)-reactive CD4^+^ T cells in peripheral blood. We demonstrate the presence of S-reactive CD4^+^ T cells in 83% of COVID-19 patients, as well as in 34% of SARS-CoV-2 seronegative healthy donors (HD), albeit at lower frequencies. Strikingly, S-reactive CD4^+^ T cells in COVID-19 patients equally targeted N-terminal and C-terminal epitopes of S whereas in HD S-reactive CD4^+^ T cells reacted almost exclusively to the C-terminal epitopes that are a) characterized by higher homology with spike glycoprotein of human endemic “common cold” coronaviruses (hCoVs), and b) contains the S2 subunit of S with the cytoplasmic peptide (CP), the fusion peptide (FP), and the transmembrane domain (TM) but not the receptor-binding domain (RBD). In contrast to S-reactive CD4^+^ T cells in HD, S-reactive CD4^+^ T cells from COVID-19 patients co-expressed CD38 and HLA-DR, indivative of their recent *in vivo* activation. Our study is the first to directly measure SARS-CoV-2-reactive T cell responses providing critical tools for large scale testing and characterization of potential cross-reactive cellular immunity to SARS-CoV-2. The presence of pre-existing SARS-CoV-2-reactive T cells in a subset of SARS-CoV-2 naïve HD is of high interest but larger scale prospective cohort studies are needed to assess whether their presence is a correlate of protection or pathology for COVID-19. Results of such studies will be key for a mechanistic understanding of the SARS-CoV-2 pandemic, adaptation of containment methods and to support vaccine development.

## Main

The COVID-19 pandemic poses an unprecedented threat to public health and the global economy. On 9^th^ April 2020, the number of confirmed COVID-19 cases worldwide surpassed 1,500,000 cases with over 90,000 COVID-19 related deaths^1^. Diagnosis of COVID-19 is routinely achieved by detection of SARS-CoV-2 RNA in nasopharyngeal swabs via PCR^5^, which works reliably in the acute phase of COVID-19^6,7^. However, limited test availability and preferential testing of symptomatic patients has likely lead to significant underestimation of infection burden and overestimation of mortality rates^8^. Whereas serological analysis of SARS-CoV-2-induced humoral immunity could reveal asymptomatic infections, it is not yet widely applied^9,10^ and complicated by the fact that coronavirus-induced antibody responses are quite variable and rather short-lived^11,12^. Coronavirus-induced cellular immunity is predicted to be more sustained, but poorly characterized so far. However, a number of T cell epitopes in coronavirus structural proteins have been predicted or identified^11,13–15^. Importantly, T helper cell responses and generation of neutralizing antibodies may be interdependent^11,16^. Studies on the SARS-CoV epidemic in 2002/03 have shown that adaptive immune responses towards spike glycoprotein were protective^11,17,18^. Hence, induction of SARS-CoV-2-specific CD4^+^ T cells is likely to be critical in the instruction of affinity maturated and potentially protective antibody responses^19^. We therefore examined the presence, frequencies and phenotypic characteristics of SARS-CoV-2 spike glycoprotein (S)-reactive T cells in COVID-19 patients compared to SARS-CoV-2 seronegative healthy donors (HD).

### Direct identification of SARS-CoV-2 spike glycoprotein-reactive CD4^+^ T cells

SARS-CoV-2 spike glycoprotein (S)-reactive CD4^+^ T cells were identified according to their expression of CD40L and 4-1BB after *in vitro* stimulation of peripheral blood mononuclear cells (PBMCs) using spike glycoprotein peptides. To this end, we designed two overlapping peptide pools (15 amino acids (aa), 11 aa overlaps) spanning the entire S that comprised different amounts of putative MHC-II epitopes according to experimental data for identified epitopes in SARS-CoV^13–15^ (Figure 1a). Peptide pool PepMix^™^ 1 (henceforth referred to as S-I) comprised overlapping peptides covering the N-terminal part (aa residues 1-643) including 21 predicted SARS-CoV MHC-II epitopes (Figure 1a, Supplementary Figure 1, Supplementary Table 1). The second peptide pool PepMix^™^ 2 (henceforth referred to as S-II) covered the C-terminal portion (amino acid residues 633-1273) including 13 predicted SARS-CoV MHC-II epitopes (Figure 1a). Peptide pool S-II exhibits a higher homology with human endemic “common cold” coronaviruses (HCoV; 229E, NL63, OC43, HKU1) with regard to the SARS-CoV MHC-II epitopes, as compared to peptide pool S-I (Figure 1a, b). The peptides of the receptor-binding domain (RBD) in the subunit S1, which represents a major target of neutralizing antibodies, are included in S-I^20,21^. For antigen-specific stimulation, PBMCs were isolated from blood samples of COVID-19 patients and HD (see Table 1 and Table 2 for patient characteristics and Supplementary Table 2 for HD characteristics) and stimulated for 16 hours with S-I and S-II peptide pools, respectively. Antigen-reactive CD4^+^ T cells were identified by co-expression of 4-1BB and CD40L, enabling the sensitive detection of S-reactive CD4^+^ T cells re-activated by TCR engagement *ex vivo*^22–24^ (Figure 2a, Supplementary Figure 2). Twelve of 18 COVID-19 (67%) patients had detectable S-I-reactive CD4^+^ T cells. Fifteen COVID-19 (83%) patients had detectable S-reactive CD4^+^ T cells against S-II peptide pool (Figure 2b, d). Most non-reactive COVID-19 patients were characterized by critical disease states (Supplementary Figure 3). Remarkably, S-reactive CD4^+^ T cells could also be detected in 23 (34%) of 68 HD, albeit at lower frequencies compared with COVID-19 patients (Figure 2e). These HD are henceforth defined as reactive healthy donors (RHD). Thereof, S-I-reactive CD4^+^ T cells could only be detected in 6 of these 23 RHD, i.e. in 8.8% of all HD. All HD were negative for IgG antibodies specific for S subunit 1 (S1) in contrast to COVID-19 patients (Figure 2f). We also ruled out early SARS-CoV-2 infection in 10 RHD by PCR standard diagnosis of nasopharyngeal swabs (data not shown).

**Table 1.** Baseline characteristics of hospitalized COVID-19 patients. *Day after onset of symptoms, ICU: intensive care unit, y: yes, n: no, m: male, f: female,

| COVID-19 patients |  |  |  |  |  |  |
| --- | --- | --- | --- | --- | --- | --- |
| ID | Severity | Gender | Age | Chief complaints at admission | ICU (y/n) | Sampling (day)* |
| P01 | mild | m | 21 | Fever, dry cough, malaise | n | 39 |
| P03 | mild | m | 45 | Fever, dry cough, runny nose | n | 25 |
| P10 | mild | f | 50 | Fever, dry cough, myalgia, cephalgia, diarrhea | n | 17 |
| P18 | mild | f | 60 | Fever, myalgia, cephalgia, runny nose | n | 21 |
| P19 | mild | m | 52 | Dry cough, cephalgia, arthralgia, nausea | n | 11 |
| P23 | mild | f | 44 | Fever | n | 5 |
| P27 | mild | f | 41 | Fever, dry cough | n | 13 |
| P06 | severe | m | 24 | Fever, dyspnea, malaise | y | 11 |
| P07 | severe | m | 33 | Fever, dry cough, dyspnea, myalgia | n | 5 |
| P11 | severe | m | 61 | Fever, dry cough, dyspnea, sore throat | y | 12 |
| P16 | severe | m | 74 | Fever, dry cough, dyspnea, malaise | y | 12 |
| P24 | severe | m | 64 | Fever, dry cough, dyspnea | y | 19 |
| P08 | critical | m | 63 | Fever, dyspnea | y | 2 |
| P12 | critical | m | 75 | Fever, dyspnea, malaise | y | 8 |
| P14 | critical | m | 81 | Fever, dyspnea | y | 8 |
| P15 | critical | m | 54 | Dry cough, dyspnea | y | 6 |
| P20 | critical | f | 53 | Dry cough, dyspnea | y | 11 |
| P21 | critical | m | 52 | Dry cough, dyspnea | y | 9 |

**Table 2.** Baseline characteristics of COVID-19 patients and healthy donors. *Day after onset of symptoms, Avg: average, m: male, f: female, ICU: intensive care unit, y: yes, n: no.

| Cohort | m:f | Age<br>Avg (range) | severity |  | ICU (y/n) | Sampling day<br>Avg (range) |
| --- | --- | --- | --- | --- | --- | --- |
| **COVID-19 patients** | 72 : 28 % | 52.6<br>(21-81 yrs) | mild | 38.9% | n 44.4% | 14.9<br>(2-39) |
|  |  |  | severe | 27.8% | y 55.6% |  |
|  |  |  | critical | 33.3% |  |  |
| **Healthy donors** | 31 : 59 % | 41.9<br>(20-64 yrs) | - | - | - | - |

**Figure 1:**
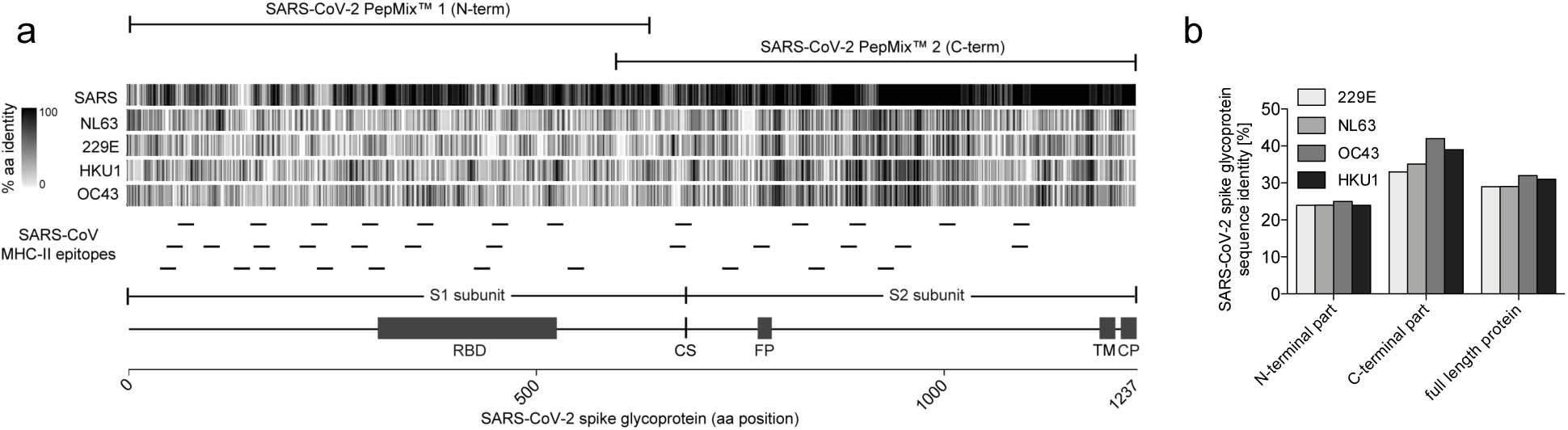
Structural domains, homology and MHC-II epitopes of the SARS-CoV-2 spike glycoprotein. **a**, SARS-CoV-2 spike glycoprotein (1237 amino acids (aa)) is separated at the cleavage site (CS) into subunit S1 harboring the receptor-binding domain (RBD) and subunit S2 containing the fusion peptide (FP), the transmembrane domain (TM) and the cytoplasmic peptide (CP). Sequence homology of spike glycoprotein of SARS-CoV-2 to SARS-CoV and HCoV strains NL63, 229E, HKU1 and OC43 was calculated as percentage of aa identity in sliding windows of 10 aa and is depicted as grey scale bars. Known SARS-CoV MHC-II epitopes are indicated as small lines below and sequences are listed in Supplementary Table 2. Homology is depicted for each reported MHC-II epitope in Supplementary Figure 1. SARS-CoV-2 PepMix™ 1 (N-term) (refered to as S-I) spans over the N-terminal and SARS-CoV-2 PepMix™ 2 (C-term) (refered to as S-II) over the C-terminal part of S. **b**, Proportion of sequence identity of the N-terminal and C-terminal parts of SARS-CoV-2 spike glycoprotein to the spike glycoproteins of HCoV strains NL63, 229E, HKU1 and OC43.

**Figure 2:**
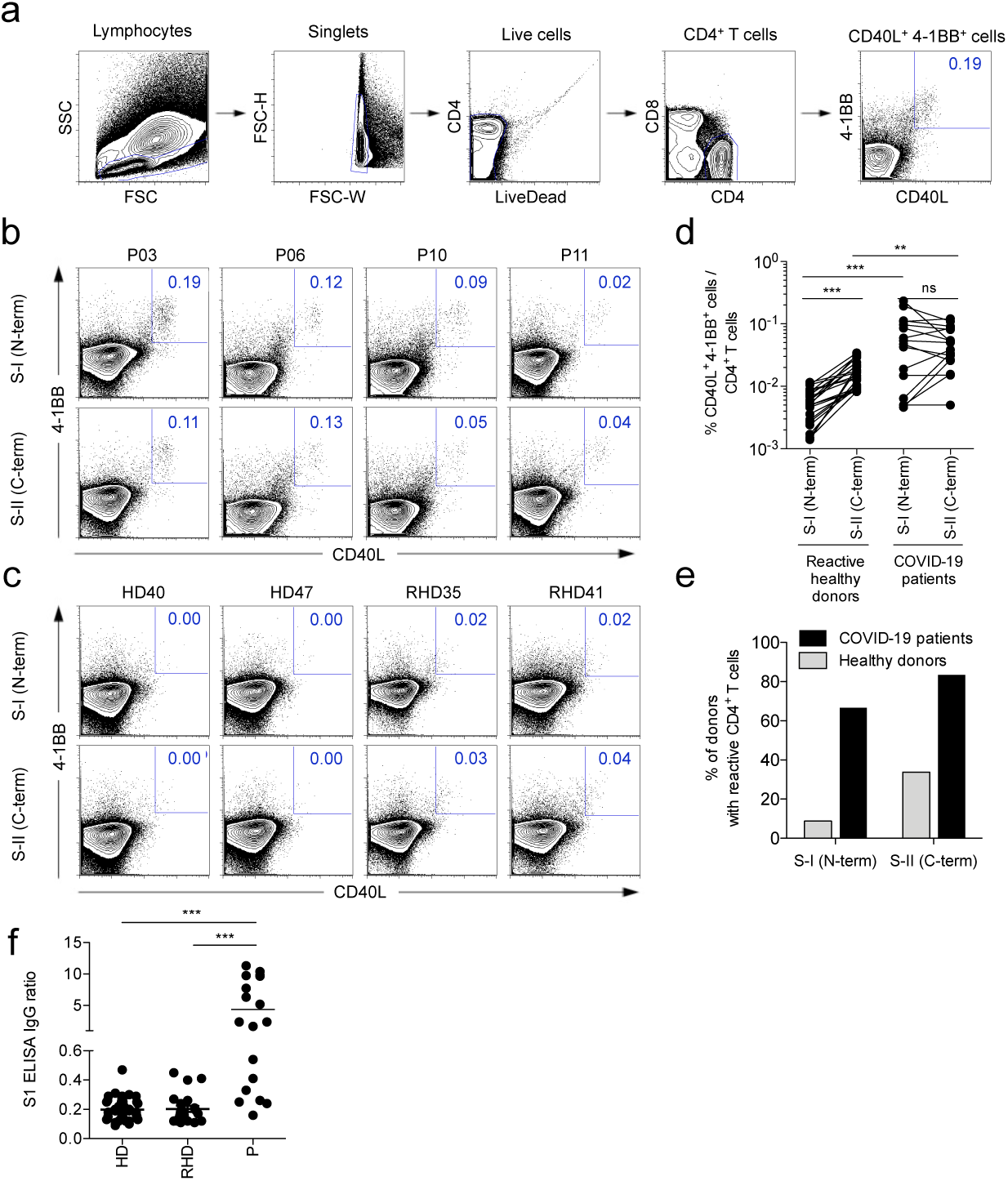
SARS-CoV-2 spike glycoprotein-reactive CD4^+^ T cells in COVID-19 patients and healthy donors. **a**, Gating strategy to detect SARS-CoV-2-reactive CD4^+^ T cells after 16 hours *in vitro* stimulation with PepMix™ SARS-CoV-2 spike glycoprotein peptide pool 1 (S-I (N-term)) and 2 (S-II (C-term)). Representative result of one COVID-19 patient is depicted. **b, c**, Representative plots displaying CD40L and 4-1BB expression on CD4^+^ T cells of COVID-19 patients (P), healthy donors (HD) and reactive healthy donors (RHD) after 16 hours *in vitro* stimulation with S-I (N-term) or S-II (C-term). Numbers indicate percent of total CD4^+^ T cells. **d**, Comparison of S-I (N-term)- or S-II (C-term)-reactive CD40L^+^4-1BB^+^ frequencies of RHD (*n*=23) and COVID-19 patients (*n*=18). **e**, Ratio of S-I (N-term)- or S-II (C-term)-reactive individuals within the cohort of COVID-19 patients and HD. **f**, Comparison of anti-spike glycoprotein subunit 1 (S1) IgG titers (ratio normalized to calibrator well) of HD (*n*=44), RHD (*n*=23) and P (*n*=18). * p<0.05, ** p<0.01, *** p<0.001 as calculated by two-tailed Mann-Whitney U test.

### Spike N-terminal versus C-terminal CD4^+^ T cell reactivity delineates COVID-19 patients from RHD

S-reactive CD4^+^ T cells in COVID-19 equally targeted both N-terminal (S-I) and C-terminal peptide pools (S-II) of S (Figure 2d). In contrast, S-reactive CD4^+^ T cells from HD exhibited a significantly stronger reactivity to the C-terminal peptide pool S-II, characterized by higher homology to spike glycoprotein of HCoV, compared to the N-terminal pool S-I (Figure 2d). The data suggest that S-reactivity among CD4^+^ T cells in SARS-CoV-2-naïve HD originated from previous immune responses against HCoV. Therefore, we additionally tested 18 of the 68 HD for the presence of antibodies specific for the four HCoV (229E, NL63, HKU1, OC43). We detected IgG antibodies against the four HCoV in all tested HD, regardless of the presence of measurable S-reactive CD4^+^ T cells (Supplementary Figure 4), showing that S- (cross-)reactive CD4^+^ T cells from HD do not correlate with antibody levels against HCoVs. This is in line with findings from other anti-viral CD4^+^ T cell responses such as in the course of yellow fever vaccination (YFV-17D). Only at very early time points after YFV-17D vaccination CD4^+^ T cell responses correlated with generation of high titers of neutralizing antibodies measured at later time points such as day 14^25^.

### Specific activation signatures of S-reactive CD4^+^ T cells in COVID-19 patients

Recent studies on SARS-CoV-2 serology have raised the question whether antibody studies are sufficient to identify individuals with mild or asymptomatic courses of COVID-19^7,10^ or whether additional, cellular signatures are needed to identify acute or past infections. Here, we assessed additional activation marker profiles on S-reactive T cells from COVID-19 patients. Expression of CD38, HLA-DR and Ki-67 has previously been shown to reliably characterize recently *in vivo* activated human T cells during acute and chronic infection^26–30^. Notably, S-reactive CD4^+^ T cells from COVID-19 patients largely expressed CD38, HLA-DR and Ki-67 (Figure 3a-d). The majority of S-reactive T cells in COVID-19 patients co-expressed CD38 and HLA-DR (Figure 3e) characteristic for effector T cell responses during acute viral infections^26,28^ whereas CD38 and Ki-67 co-expression was more variable (Figure 3f). By contrast, S-reactive CD4^+^ T cells from HD did not express CD38, HLA-DR and Ki-67, or only at low frequencies (Figure 3b-f), and co-expression was not observed (Figure 3e, f). In COVID-19 patients, considerable proportions of peripheral CD4^+^ and CD8^+^ T cells co-expressed CD38 and HLA-DR (data not shown), which, however, could not be re-activated with our S peptide pools *in vitro*. These findings are in line with results of a recent study showing refractory T cell signatures in COVID-19 patients^31^ and also a proportion of these CD38^+^HLADR^+^ CD4^+^ T cells may target other structural proteins of SARS-CoV-2. We also show that the presence of S-reactive CD4^+^ T cells and in particular of CD38 expressing cells among S-reactive CD4^+^ T cells exhibited a high variability among patients in the course of COVID-19 disease (Figure 3g,h).

**Figure 3:**
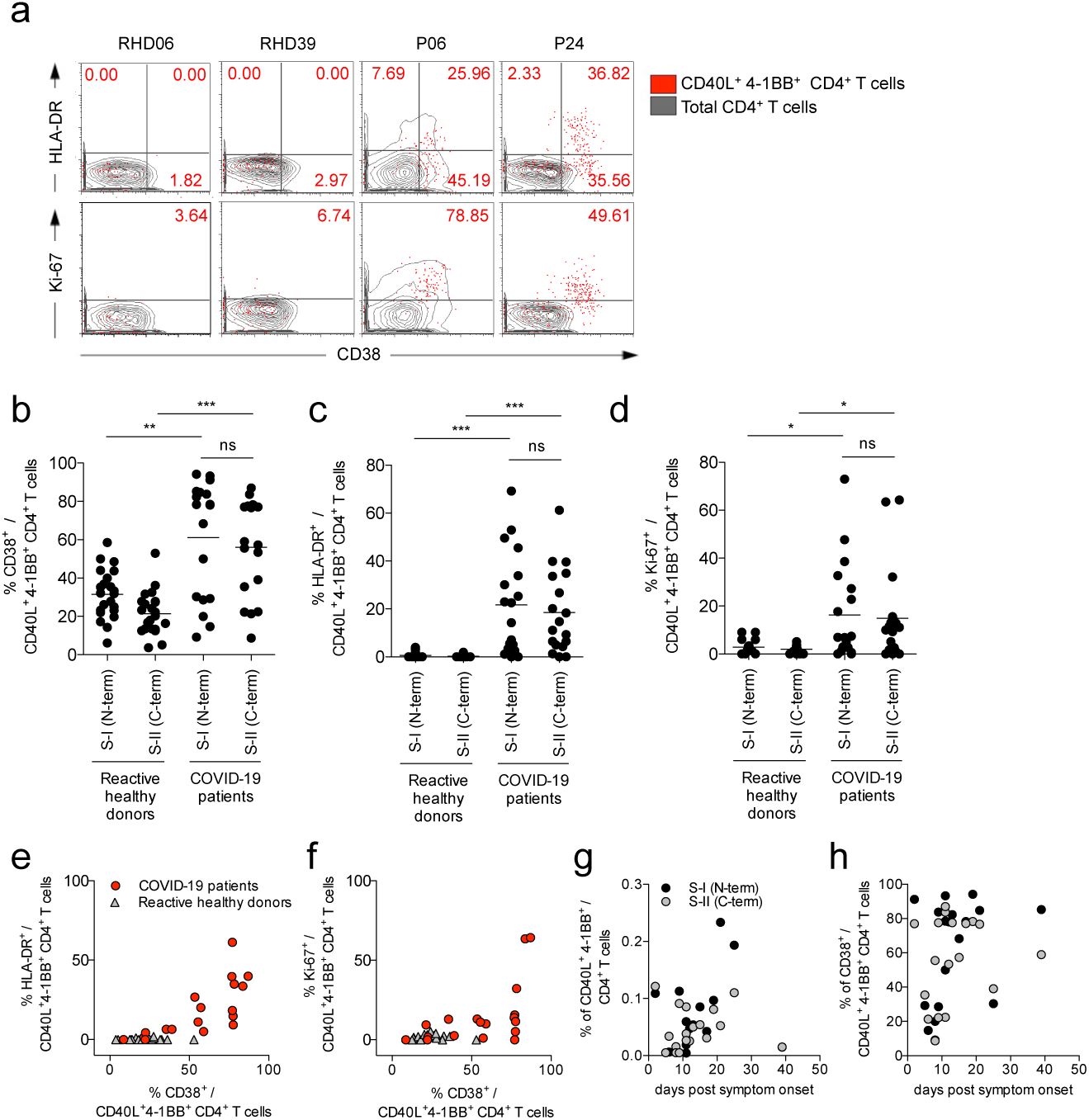
CD38, HLA-DR and Ki-67 expression of SARS-CoV-2 S-I and S-II-reactive CD4^+^ T cells discriminates SARS-CoV-2 patients from reactive healthy donors. **a**, Representative examples of HLA-DR or Ki-67 against CD38 expression on S-II (C-term)-reactive CD4^+^ T cells (red dots) compared to total CD4^+^ T cells (grey contours) in RHD and COVID-19 patients. **b-d**, Comparison of frequencies of CD38^+^, HLA-DR^+^ and Ki67^+^ cells among S-I (N-term)- and S-II (C-term)-reactive CD4^+^ T cells in RHD (**b**,**c**, *n*=23; **d**, *n*=17) and COVID-19 patients (*n*=18). * p<0.05, ** p<0.01, *** p<0.001 as calculated by two-tailed Mann-Whitney U test. **e, f,** Co-expression of HLA-DR^+^ or Ki-67^+^ and CD38^+^ among S-II (C-term)-reactive CD4^+^ T cells from RHD (**e**, *n*=23; **f**, *n*=17) and COVID-19 patients (*n*=18). **g, h**, Frequencies of S-I (N-term)- and S-II (C-term)-reactive CD40L^+^ 4-1BB^+^ CD4^+^ T cells or CD38^+^ among S-II (C-term)-reactive cells of COVID-19 patients (*n*=18) plotted over time (days post symptom onset).

## Discussion

Our study demonstrates the presence of S-reactive CD4^+^ T cells in COVID-19 patients, and in a subset of SARS-CoV-2 seronegative HD. In light of the very recent emergence of SARS-CoV-2, our data raise the intriguing possibility that pre-existing S-reactive T cells in a subset of SARS-CoV-2 seronegative HD represent cross-reactive clones raised against S-proteins, probably acquired as a result of previous exposure to HCoV. Endemic HCoV account for about 20% of “common cold” upper respiratory tract infections in humans. HCoV infections are ubiquitous, but they display a winter seasonality in temperate regions^32–34^. Based on epidemiological data indicating an average of two episodes of “common cold” per year in the adult population, it may be extrapolated that the average adult contracts a HCoV infection on average every two to three years. Protective antibodies may wane in the interim but cellular immunity could remain^15,35^. Although the overall amino acid sequence homology of spike glycoproteins is relatively low among HCoV, there is an overlap of MHC-II epitopes located especially in the C-terminal domain of the here used peptide pools (Figure 1a, Supplementary Figure 1). This may explain the preferential reactivity of CD4^+^ T cells to the C-terminal domain in one third of SARS-CoV-2 seronegative HD.

The biological role of pre-existing SARS-CoV-2 S-cross-reactive CD4^+^ T cells in 34% of HD remains unclear for now. However, these cells may represent the key to understanding the vastly divergent manifestations of SARS-CoV-2 disease courses, and particularly the suspected high rate of asymptomatic infections in children and young adults assuming that these S-cross-reactive CD4^+^ T cells have a protective role in SARS-CoV-2 infection. Since children and young adults have on average more frequent social contacts than the elderly, one might expect a higher transmission rate and HCoV prevalence in the former. This assumption would need to be investigated in future longitudinal studies assessing the presence of pre-existing SARS-CoV-2-cross-reactive CD4^+^ T cells and their impact on the susceptibility to SARS-CoV-2 infcetion and age-realted clinical outcomes of COVID-19.

SARS-CoV neutralizing antibodies are associated with convalescence, and they have been detected 12 months after disease^11^. However, the durability of neutralizing antibody responses against SARS-CoV-2 currently remains unknown. Antibodies against HCoV can wane within months after infection, although HCoV re-infection is accompanied by low-level and short-lived virus shedding with only mild symptoms of short duration^12^ pointing towards residual immunity. Cellular immunity has not yet been studied in this context. The extent to which and how SARS-CoV-2 specific humoral or cellular immunity mediates durable protection against reinfection is unknown but will be one of the critically important fields of research in the coming months. It has been demonstrated in mouse models that CD4^+^ as well as CD8^+^ T cell responses directed against structural proteins such as spike or nucleocapsid protein of SARS-CoV critically contribute to viral clearance^17,36,37^.

To our knowledge, this study represents the first report of cellular SARS-CoV-2-cross-reactivity in human T cells. Our results provide a decisive rationale to initiate worldwide prospective studies to assess the contribution of pre-existing SARS-CoV-2-cross-reactive immunity (potential regional differences) to clinical outcomes of SARS-CoV-2-infections. Together with currently introduced, novel serology tests, the data generated by such studies may critically inform evidence-based risk evaluation, patient monitoring, adaptation of containment methods, and last but not least, vaccine development.

## Data Availability

All data referred two the manuscript ist available.

## Acknowledgements

We thank Ulf Klein (Leeds, UK) und Hans-Peter Herzel (Berlin) for critical discussion. This work was supported by the German Research Foundation (KFO339 to J.B and F.F., SFB-TR84 projects A4, B6 to S.H., B8 to M.M., C6 to M.W., C8, C10 to L.E.S., C9 to M.W, N.S., and by the German Federal Ministry of Education and Research (BMBF-RAPID to S.H., C.D., and CAPSyS to M.W., N.S.).

## Materials and Methods

### Study subjects

This study was approved by the Institutional Review board of the Charité (EA2/066/20). After providing written informed consent, 68 healthy donors (Supplementary Table 1) and 18 COVID-19 patients (Table 1) were included in the study. COVID-19 patients tested positive for SARS-CoV-2 RNA in nasopharyngeal swabs were recruited at Charité Campus Virchowklinikum, Berlin between March 1^st^ and April, 2^nd^ 2020.

### Serology

Anti-SARS-CoV-2 IgG ELISA was performed using a commercial kit (EUROIMMUN Medizinische Labordiagnostika AG) as described and validated before^38^. Recombinant immunofluorescence assays (rIFA) to determine IgG titers against HCoV was done by using VeroB4 cells expressing cloned recombinant coronavirus spike proteins from HCoV-229E, HCoV-NL63, HCoV-OC43, HCoV-HKU1 as described in Corman et al.^39^.

### Cell isolation and stimulation

Peripheral blood mononuclear cells (PBMC) were isolated from heparinized whole blood by gradient density centrifugation according to manufacturer’s instructions (Leucosep tubes, Greiner; Biocoll, Bio&SELL). Stimulation was conducted with 5×10^6^ PBMC in RPMI 1640 medium (Gibco) supplemented with 10% heat inactivated AB serum (Pan Biotech), 100 U/ml penicillin (Biochrom), 0.1 mg/ml streptomycin (Biochrom) and PepMix™ SARS-CoV-2 spike glycoprotein (JPT) peptide pool 1 or 2 in the presence of 1 µg/ml purified anti-CD28 (clone CD28.2, BD Biosciences). The PepMix™ SARS-CoV-2 spike glycoprotein pool 1 covering the N-terminal amino acid (aa) residues 1-643 (abbreviated to “S-I (N-term)”) contained 158 15-mers overlapping by 11 aa. PepMix™ SARS-CoV-2 spike glycoprotein pool 2 covered the C-terminal aa residues 633-1273 (abbreviated to “S-II (C-term)”) containing 156 15-mers overlapping by 11 aa and one 17-mer at the C-terminus, i.e. 157 peptides in total. Both peptide pools were used at 1 µg/ml per peptide, respectively. Further details on the peptide pools and predicted MHC-II epitopes are given in Figure 1, Supplementary Figure 1 and Supplementary Table 2. Stimulation controls were performed with equal concentrations of DMSO in PBS (unstimulated) or 1.5 mg SEB/1.0 mg TSST1 (Sigma-Aldrich) and PepMix™ HCMVA (pp65) (>90%) (JPT) in the presence of 1 µg/ml purified anti-CD28 (clone CD28.2, BD Biosciences) as positive controls, respectively. Incubation was performed at 37°C, 5% CO2 for 16 h with 10 µg/ml brefeldin A (Sigma-Aldrich) added after 2 h.

### Flow Cytometry

Stimulation was stopped by incubation in 20 mM EDTA for 5 min and surface staining conducted for 15 min with the following fluorochrome conjugated antibodies titrated to their optimal concentrations: CD38-PE-Vio770 (clone REA671, Miltenyi), CD69-APC-Cy7 (FN50, Biolegend), HLAD-DR-VioGreen (REA805, Miltenyi), CD4-BrilliantViolet605 (RPA-T4, Biolegend), CD8-PerCP (SK1, Biolegend) with 1 mg/ml Beriglobin (CSL Behring) added prior to the staining. For exclusion of dead cells, Zombie Yellow fixable viability staining (Biolegend) was added for the last 10 min of incubation. Fixation and permeabilization were performed with eBioscience™ FoxP3 fixation and PermBuffer (Invitrogen) according to the manufacturer’s protocol and intracellular staining carried out for 30 min in the dark at room temperature with Beriglobin added prior to intracellular staining with 4-1BB-PE (clone 4B4-1, BD), CD40L-APC (5C8, Miltenyi) and Ki-67-AlexaFluor488 (B56, BD). Samples were measured on a MACSQuant^®^ Analyzer 16. Instrument performance was monitored daily with Rainbow Calibration Particles (BD).

### Data analysis and statistics

Flow cytometry data were analyzed with FlowJo 9.9.6 (FlowJo LLC). Prism 5 (GraphPad Inc.) was used for plotting and statistical analysis. Non-parametric testing was used to compare cell frequencies and antibody titers between groups (two-tailed Mann-Whitney U test). *n* indicates the number of donors.

## References

1. Dong, E., Du, H. & Gardner, L. COVID-19 in real time. Lancet Infect. Dis. 3099, 19–20 (2020).

2. Wang, D. et al.. Clinical Characteristics of 138 Hospitalized Patients with 2019 Novel Coronavirus-Infected Pneumonia in Wuhan, China. JAMA - J. Am. Med. Assoc. (2020). doi:10.1001/jama.2020.1585

3. Channappanavar, R., Zhao, J. & Perlman, S. T cell-mediated immune response to respiratory coronaviruses. Immunologic Research (2014). doi:10.1007/s12026-014-8534-z

4. Li, C. K. et al.. T Cell Responses to Whole SARS Coronavirus in Humans. J. Immunol. 181, 5490–5500 (2008).

5. Corman, V. M. et al.. Detection of 2019 novel coronavirus (2019-nCoV) by real-time RT-PCR. Euro Surveill. (2020). doi:10.2807/1560-7917.ES.2020.25.3.2000045

6. Loeffelholz, M. J. & Tang, Y.-W. Laboratory diagnosis of emerging human coronavirus infections – the state of the art. Emerg. Microbes Infect. 9, 747–756 (2020).

7. Wölfel, R. et al.. Virological assessment of hospitalized patients with COVID-2019. Nature 1–10 (2020). doi:10.1038/s41586-020-2196-x

8. Wu, J. T., Leung, K. & Leung, G. M. Nowcasting and forecasting the potential domestic and international spread of the 2019-nCoV outbreak originating in Wuhan, China: a modelling study. Lancet 395, 689–697 (2020).

9. Amanat, F. et al.. A serological assay to detect SARS-CoV-2 seroconversion in humans. medRxiv 2020.03.17.20037713 (2020). doi:10.1101/2020.03.17.20037713

10. Okba, N. M. A. et al.. SARS-CoV-2 specific antibody responses in COVID-19 patients. medRxiv 2020.03.18.20038059 (2020). doi:10.1101/2020.03.18.20038059

11. Li, C. K. et al.. T Cell Responses to Whole SARS Coronavirus in Humans. J. Immunol. 181, 5490–5500 (2008).

12. Callow, K. A., Parry, H. F., Sergeant, M. & Tyrrell, D. A. J. The time course of the immune response to experimental coronavirus infection of man Nasal washings. 435–446 (1990).

13. Libraty, D. H., O’Neil, K. M., Baker, L. M., Acosta, L. P. & Olveda, R. M. Human CD4+ Memory T-lymphocyte Responses to SARS Coronavirus Infection. Virology 368, 317–321 (2007).

14. Yang, J. et al.. Searching immunodominant epitopes prior to epidemic: HLA class II-restricted SARS-CoV spike protein epitopes in unexposed individuals. Int. Immunol. 21, 63—71 (2009).

15. Ng, O. et al.. Memory T cell responses targeting the SARS coronavirus persist up to 11 years post-infection. Vaccine 34, 2008–2014 (2016).

16. Mitchison, N. A. T-cell–B-cell cooperation. 4, 1599–1601 (2004).

17. Yang, Z. et al.. A DNA vaccine induces SARS coronavirus neutralization and protective immunity in mice. Nature 428, 561–564 (2004).

18. Zhu, Z. et al.. Potent cross-reactive neutralization of SARS coronavirus isolates by human monoclonal antibodies. Proc. Natl. Acad. Sci. 104, 12123–12128 (2007).

19. Ju, B. et al.. Potent human neutralizing antibodies elicited by SARS-CoV-2 infection. bioRxiv 2020.03.21.990770 (2020). doi:10.1101/2020.03.21.990770

20. Meng, T. et al.. The insert sequence in SARS-CoV-2 enhances spike protein cleavage by TMPRSS. (2020).

21. Walls, A. C. et al.. Structure, Function, and Antigenicity of the SARS-CoV-2 Spike Glycoprotein. Cell (2020). doi:10.1016/j.cell.2020.02.058

22. Frentsch, M. et al.. Direct access to CD4+ T cells specific for defined antigens according to CD154 expression. Nat. Med. 11, 1118–1124 (2005).

23. Sattler, A. et al.. Cytokine-induced human IFN-γ–secreting effector-memory Th cells in chronic autoimmune inflammation. Blood 113, 1948–1956 (2009).

24. Schoenbrunn, A. et al.. A Converse 4-1BB and CD40 Ligand Expression Pattern Delineates Activated Regulatory T Cells (Treg) and Conventional T Cells Enabling Direct Isolation of Alloantigen-Reactive Natural Foxp3+ Treg. J. Immunol. 189, 5985–5994 (2012).

25. Kohler, S. et al.. The early cellular signatures of protective immunity induced by live viral vaccination. Eur. J. Immunol. (2012). doi:10.1002/eji.201142306

26. Appay, V. et al.. Memory CD8+ T cells vary in differentiation phenotype in different persistent virus infections. Nat. Med. (2002). doi:10.1038/nm0402-379

27. Blom, K. et al.. Temporal Dynamics of the Primary Human T Cell Response to Yellow Fever Virus 17D As It Matures from an Effector- to a Memory-Type Response. J. Immunol. (2013). doi:10.4049/jimmunol.1202234

28. Callan, M. F. C. et al.. Direct visualization of antigen-specific CD8+ T cells during the primary immune response to Epstein-Barr virus in vivo. J. Exp. Med. 187, 1395–1402 (1998).

29. Miller, J. D. et al.. Human Effector and Memory CD8+ T Cell Responses to Smallpox and Yellow Fever Vaccines. Immunity (2008). doi:10.1016/j.immuni.2008.02.020

30. Schulz, A. R. et al.. Low Thymic Activity and Dendritic Cell Numbers Are Associated with the Immune Response to Primary Viral Infection in Elderly Humans. J. Immunol. (2015). doi:10.4049/jimmunol.1500598

31. Diao, B. et al.. Reduction and Functional Exhaustion of T Cells in Patients with Coronavirus Disease 2019 (COVID-19). medRxiv (2020). doi:https://doi.org/10.1101/2020.02.18.20024364.

32. Tyrrell, D. A. J. Common colds and Related Diseases. (1965).

33. Lidwell, O. M. & Williams, R. E. The epidemiology of the common cold. I. J. Hyg. (Lond). 59, 309–319 (1961).

34. Gaunt, E. R., Hardie, A., Claas, E. C. J., Simmonds, P. & Templeton, K. E. Epidemiology and clinical presentations of the four human coronaviruses 229E, HKU1, NL63, and OC43 detected over 3 years using a novel multiplex real-time PCR method. J. Clin. Microbiol. 48, 2940–2947 (2010).

35. Callow, K. A., Parry, H. F., Sergeant, M. & Tyrrell, D. A. The time course of the immune response to experimental coronavirus infection of man. Epidemiol. Infect. 105, 435–446 (1990).

36. Channappanavar, R., Fett, C., Zhao, J., Meyerholz, D. K. & Perlman, S. Virus-Specific Memory CD8 T Cells Provide Substantial Protection from Lethal Severe Acute Respiratory Syndrome Coronavirus Infection. J. Virol. 88, 11034–11044 (2014).

37. Wang, B. et al.. Identification of an HLA-A*0201-restricted CD8+ T-cell epitope SSp-1 of SARS-CoV spike protein. Blood 104, 200–206 (2004).

38. Okba, N. M. A. et al.. Severe Acute Respiratory Syndrome Coronavirus 2−Specific Antibody Responses in Coronavirus Disease 2019 Patients. Emerg. Infect. Dis. J. 26, 2020.03.18.20038059 (2020).

39. Corman, V. M. et al.. Assays for laboratory confirmation of novel human coronavirus (HCOV-EMC) infections. Eurosurveillance (2012). doi:10.2807/ese.17.49.20334-en

